# Mass Homicide by Firearm in Canada: Effects of Legislation

**DOI:** 10.1101/2022.03.24.22272877

**Authors:** Caillin Langmann

## Abstract

Canada implemented a series of laws regulating firearms including background checks and licensing, references, psychological questionnaires, prohibition of paramilitary style rifles, and magazine capacity restrictions in order to decrease the incidences and deaths from mass shootings. The associated effects of these laws were examined over the years 1974 to 2020.

A model was constructed using difference-in-differences analysis of firearms and non-firearms mass homicide incidences and death rates. Mass homicides were defined as a homicide due to one event involving three or more deaths.

Incidence rates of mass homicide by firearm were found to be 0.11 (95%CI 0.08, 0.14) per million compared to a non-firearm mass homicide rate of 0.12 (95% CI 0.10, 0.15) per million. Mass homicide death rates by firearm were found to be 0.39 (95% CI 0.29, 0.49) per million compared to a non-firearm mass homicide rate of 0.47 (95% CI 0.34, 0.61) per million. Overall, there is a gradual declining trend in the incidence of mass homicide by firearm (IRR 0.97 (95% CI 0.96, 0.98)) and by non-firearm (IRR 0.97 (95% CI 0.97, 0.98)). The decline in mass homicide death rate by firearm and non-firearm is IRR 0.96 (95% CI 0.95, 0.97), and IRR 0.97 (95% CI 0.96, 0.98) respectively.

No associated decrease in mass homicide incidence rates or death rates with firearms legislation was found after the implementation of background checks and prohibition of full auto firearms in 1980, by the implementation of references and psychological questionnaires in 1994, by the restriction of magazine capacity in 1994, the prohibition of paramilitary rifles in 1994, or licensing in 2001.

## Introduction

In April, 2020, a mass homicide by firearm occurred in Nova Scotia, stimulating the Government of Canada to respond by prohibiting several types of firearms including the ones used in the attack. This is not the first time the Government of Canada has responded with regulatory efforts following a mass homicide, after the Polytechnique Institute massacre in 1989, a series of legislative efforts were produced to prevent firearm deaths as well as specifically mass homicide.

The definition of mass homicide is based on somewhat arbitrary criteria, requiring four or more fatalities in some studies, three or more in others, while some accept a certain number of injuries in proxy (1). Regardless of the definition, the Government of Canada has produced firearm legislation and regulations with the intent of reducing the number of fatalities in a mass homicide by reducing the lethality and availability of firearms (2, 3). In 1978 fully automatic firearms were prohibited and background checks to acquire long guns (rifles and shotguns) were implemented near the end of 1979. Note that a permit or later a licence was required to obtain and possess handguns during the entire years studied and has been in place since 1932. In 1991 legislation was passed and came into force between 1992 and 1994 that prohibited large capacity magazines, prohibited weapons converted from fully automatic, and required two references and expanded background checks including a mental health psychological questionnaire. As well, most paramilitary rifles were prohibited, with some restricted to gun ranges only. Overall, about 550,000 firearms were prohibited at that time (4). In 2001 regulations required a license to acquire and possess firearms with renewal every five years after a background and reference check. While several studies controlling for contributing variables have shown that the associated effects of legislation with firearms deaths appear to have been equivocal, no specific examination of the association with mass homicide in Canada has been produced (5-10). Despite several high-profile events, the incidence of mass homicide in Canada has also not been studied.

In this study, the trends in incidents and deaths of firearm associated mass homicide were examined in Canada over the years 1974 to 2020. Since many studies of legislative intervention potentially suffer from errors due to confounding variables, the impact of Canadian legislation was assessed using a difference-in-differences (DiD) approach, a method that can mitigate the potential effects of confounders (11, 12). This is the first study to look at specific points of firearms legislation and regulations and the association with mass homicide by firearm in Canada.

## Materials and Methods

The number of different definitions of mass homicide in the literature presented a challenge for designing a reasonable search criteria for this study (1). A previous request to Statistics Canada for incidents including 4 or more fatalities resulted in many years where there were zero incidents and would be difficult to analyze with statistical methods. In addition, it was possible that a high bar would limit the applicability of the results as there are possibly incidents where a significant number of victims are injured as would be suggested by three fatalities. For example, the Moncton shooting in New Brunswick in 2014 resulted in “only” three people killed while two more were injured. It seems arbitrary to exclude this serious incident from analysis and therefore a cut-off of three or more deaths per incident was accepted. An excellent review of the difficulties in arriving at a conclusive definition is covered by Greene-Colozzi and Silva 2020 (1).

Mortality data was obtained from a request submitted to Statistics Canada. Specifically, a search for the number of incidents of homicide involving three or more fatalities by weapon type over the years 1974 to 2020. Archival issues make it difficult and expensive to obtain data prior to 1974. Disaggregation by the number of deaths associated with these incidents as well as by sex of victim was obtained. Homicide by firearm included rifle, shotgun, modified firearms, handgun, fully automatic firearm, and unknown type of firearm(s). Other methods of homicide were also obtained. Homicides were counted by the year submitted to Statistics Canada. Unfortunately, due to the differing methods of collection the specific type of firearm used could only be provided with any accuracy after 1991. Population data from the years 1974 to 2020 were obtained from Statistics Canada CANSIM table 051-0001.

The study was constructed with the null hypothesis that firearms legislation and regulations implemented in 1980, 1994, and 2001 were not associated with reductions in the incidence and associated deaths due to mass homicide by firearms. A Difference in differences (DiD) technique was used to construct a quasi-experimental time series analysis to compare a control group to a treatment group exposed to the interventions. The benefit of using this model is that it mitigates the effects of external confounders and potential selection bias involved in choosing independent variables to include in regression. Two dependent variables were investigated, incidence of mass homicide and associated deaths.

A Generalized DiD model was constructed to allow for the relaxation of the parallel trends assumption, in this study a model was constructed including terms to account for differing trends prior to legislation in the control and treatment group as well as changes in trends after legislation in each group (11, 12). Observational quasi-experimental designs are also unable to control for crossover from one group to another, in this case while it was expected that firearms legislation would not directly have an effect on mass homicide by other methods, it would potentially be the case that people who were unable to use firearms for homicide would be forced to choose another method and thus “crossover” into the respective non firearms groups. Constructing a model that includes all pre and post trends can allow for an accounting of the crossover.

The Generalized DiD model was constructed with variables for year (xi1), cause of death: firearms or other (xi2), and a variable to account for whether legislation was in effect (xi3). The model utilized the variable “year (xi1)” as a term to construct a linear time trend, with interaction terms to allow for different time trends by the cause of death. To account for whether there is a variation in changes in each category, an interaction between the step term, legislation in effect, and cause of death was included. To allow for a common effect on the trend, an interaction between year and the step term was included. Finally, a 3-way interaction between year, cause of death, and legislation was included and is the difference in difference term that represented the additional effect of legislation. The population of each cohort at that year, ni, was used in the model as an offset to ensure changes in population were accounted for. The equation is written as follows:

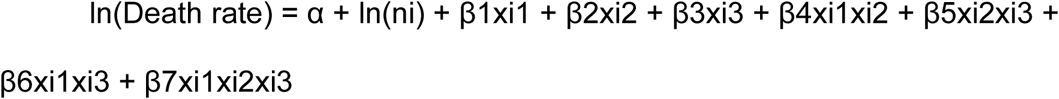

The intercept term is indicated by α. The coefficient β1 measures the time trend in non firearm mortality before the implementation of legislation, β2 measures the rate ratio of mortality in firearm vs. non firearm mortality at the starting year (1981), β3 measures the level change in non firearm mortality after the implementation of legislation, β4 measures the difference in trend for firearms relative to non firearms before the implementation of legislation, and β5 measures the level change in firearm mortality after the implementation of legislation relative to non firearm mortality. The coefficient, β6, measures the change in trend in non firearm mortality after the implementation of legislation. Finally, the 3-way interaction coefficient, β7, measures the additional change in trend in firearm mortality relative to non firearm mortality after legislation, is known as the difference in difference coefficient, and if significant it indicates that the effect of legislation on time trends differed between the non firearm and firearm categories. This 3-way term, β7, is the specific measure of the impact of the intervention.

Linear combinations of the coefficients were calculated to give an estimate of the annual change in the mortality rate before and after legislation. The DiD coefficient was expressed as an incidence rate ratio (IRR) of the additional rate of firearm mortality post legislation (12).

Analysis was conducted over the years 1974 to 2020. Impacts were set at years 1980, 1994, and 2001 to test for the effects of the implementation of each legislation. Years prior to impacts were coded as 0 and years post impact were coded as 1. The year 1980 was chosen as the implementation of background and reference checks and the prohibition of fully automatic firearms occurred at that time. The year 1994 was selected as regulations occurred that prohibited large capacity magazines, prohibited weapons converted from fully automatic, and regulations required two references and expanded background checks including a mental health psychological questionnaire prior to acquisition. As well, most paramilitary rifles were prohibited. Finally, 2001 was chosen as it was the year that all firearms owners were required to have a firearms license rather than just a certificate to acquire firearms.

Statistical analysis of incidents used negative binomial regression with errors estimated by clustered robust errors using the variable year for observation level random effects in Stata/IC version 14 (StataCorp LP, College Station, Texas). Estimates of deaths were produced using zero-inflated negative binomial regression with year and incident in the inflated regression to account for the alternative causes of zero yearly deaths. The acceptance level of statistical significance used in the analysis was a p value less than 0.05 and 95% confidence intervals (CI).

## Results

Fig 1A, Fig 1B, Fig 1C, and Fig 1D respectively display the total incidence of mass homicide per 1,000,000, total deaths by mass homicide per 1,000,000, and male and female deaths per 1,000,000 over the years 1974 to 2020. Visual inspection revealed that the incidence of mass homicide and death rates appear similar between the firearms and non-firearms cohorts. Overall, there is a gradual declining trend in the incidence of mass homicide by firearm, IRR 0.97 (95% CI 0.96, 0.98) and by non-firearm, IRR 0.97 (95% CI 0.97, 0.98) per year. The decline in mass homicide death rate by firearm and non-firearm is also notable, IRR 0.96 (95% CI 0.95, 0.97), and IRR 0.97 (95% CI 0.96, 0.98) respectively.

**Figure 1A.**
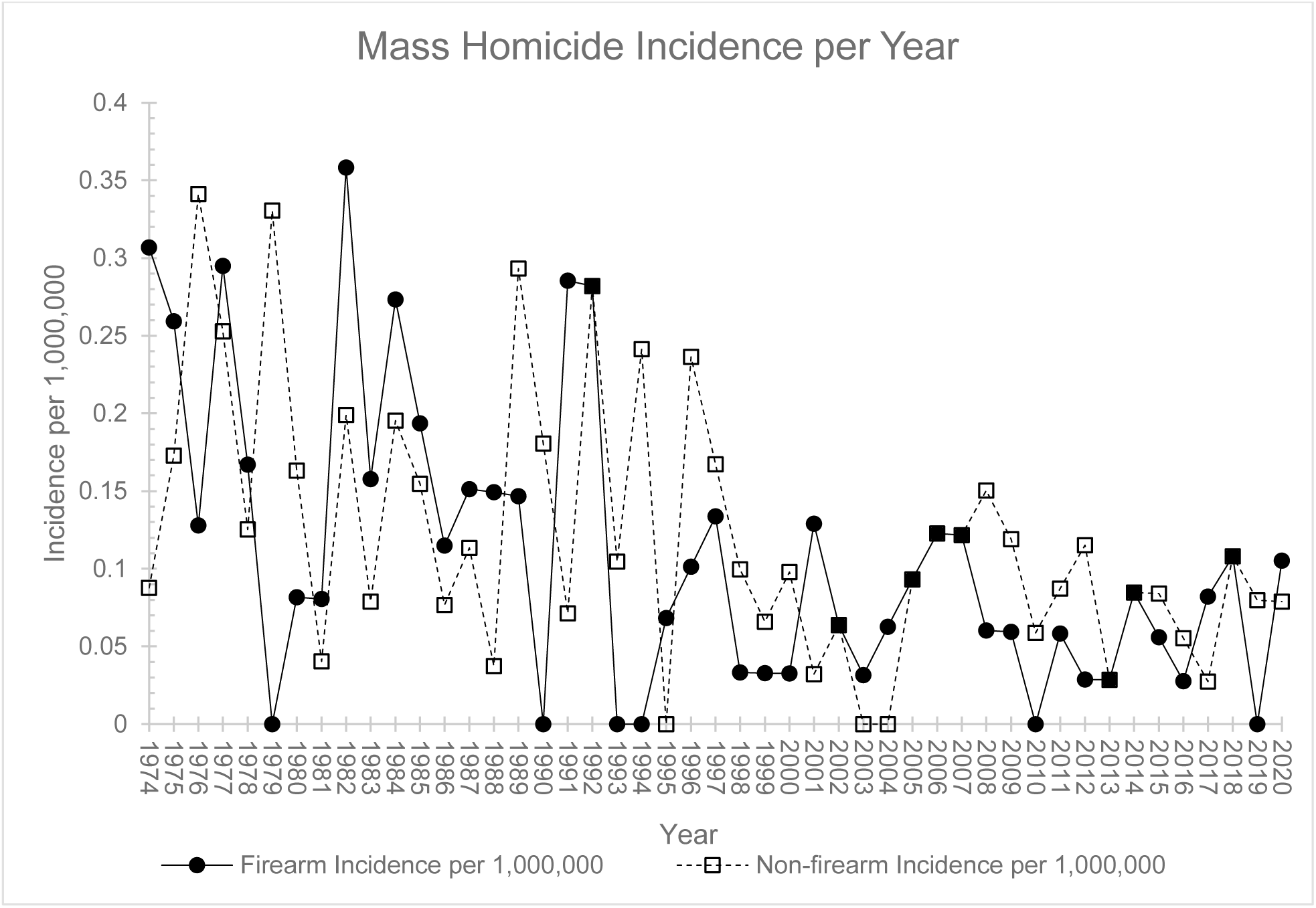

**Figure 1B.**
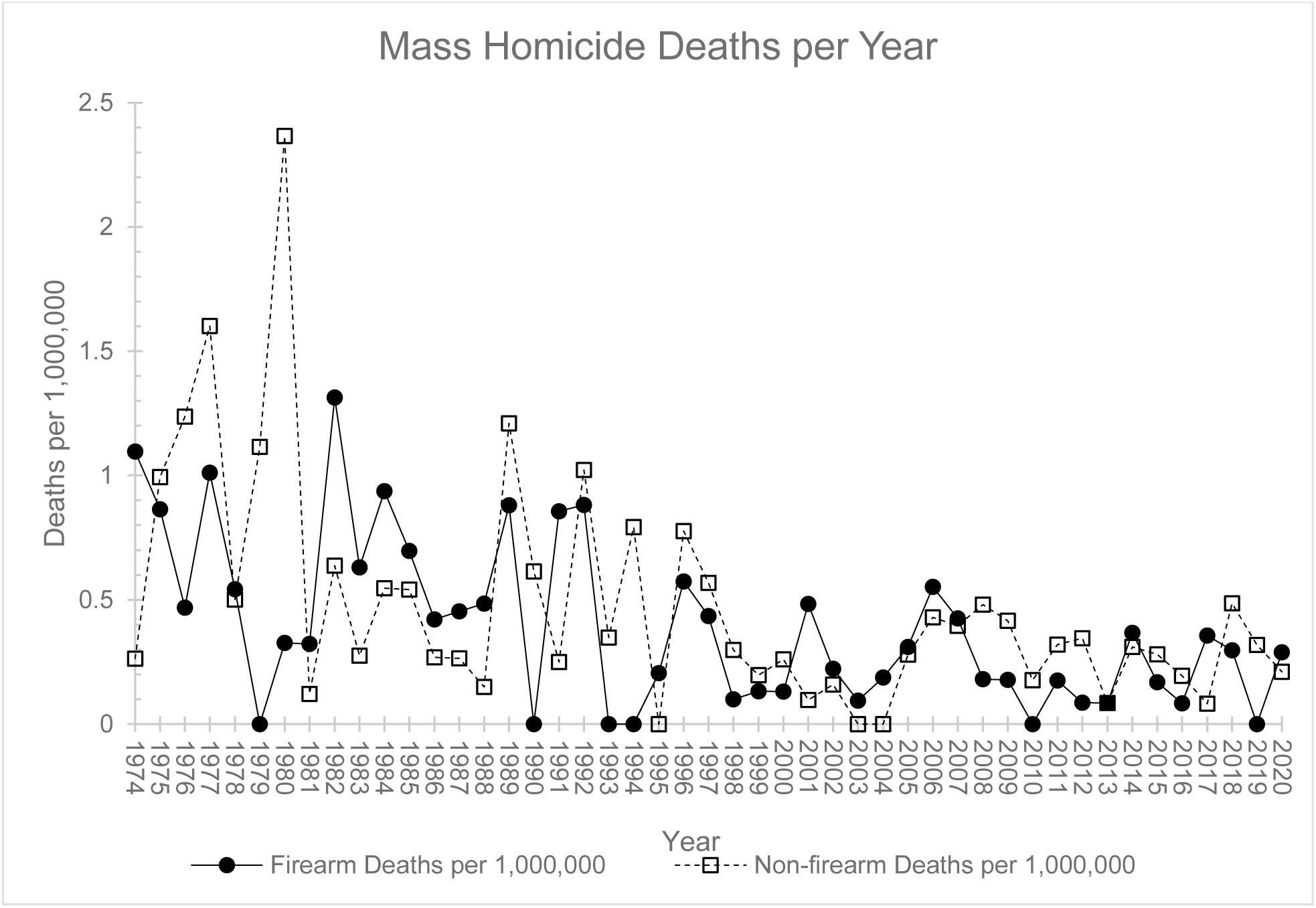

**Figure 1C.**
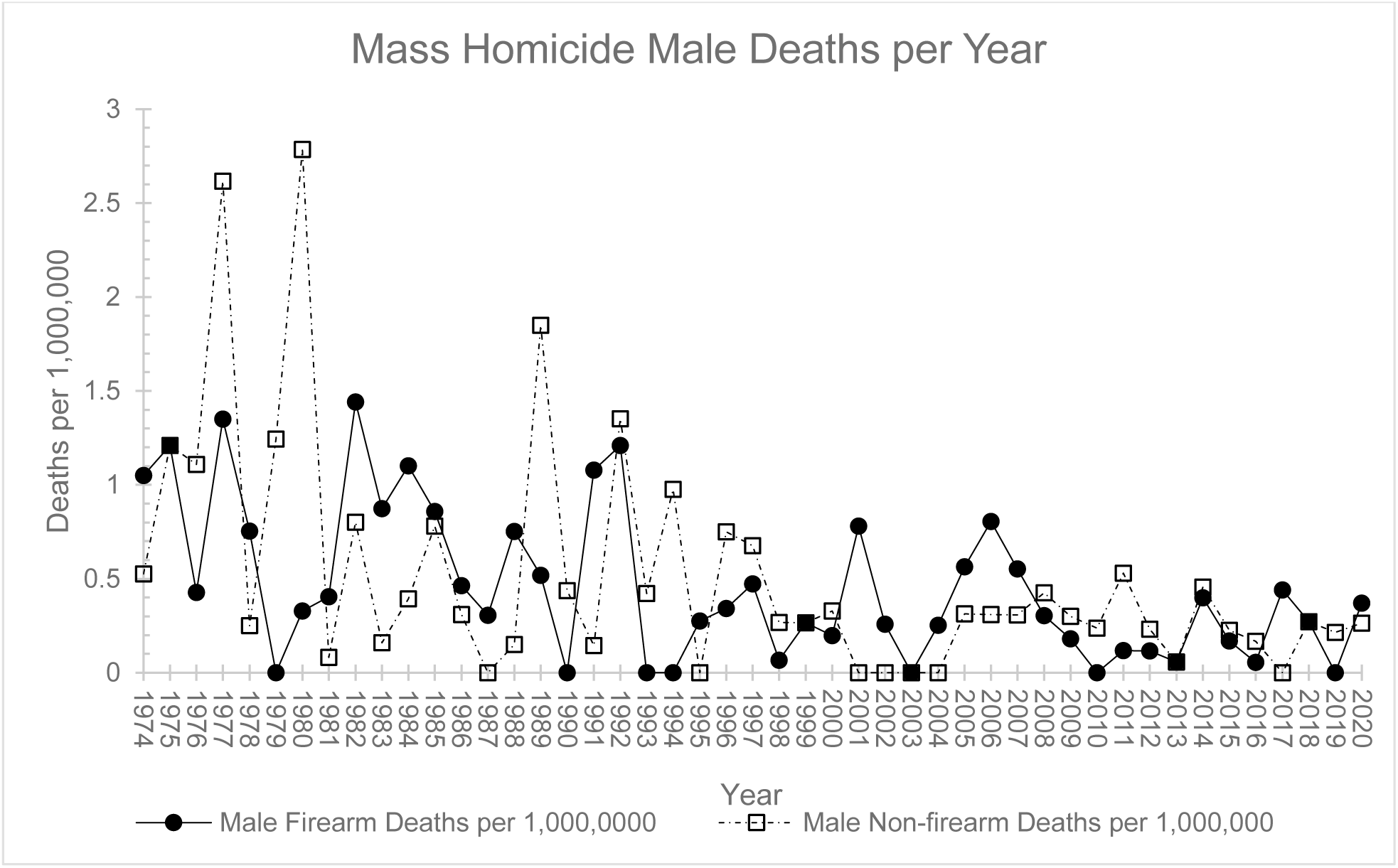

**Figure 1D.**
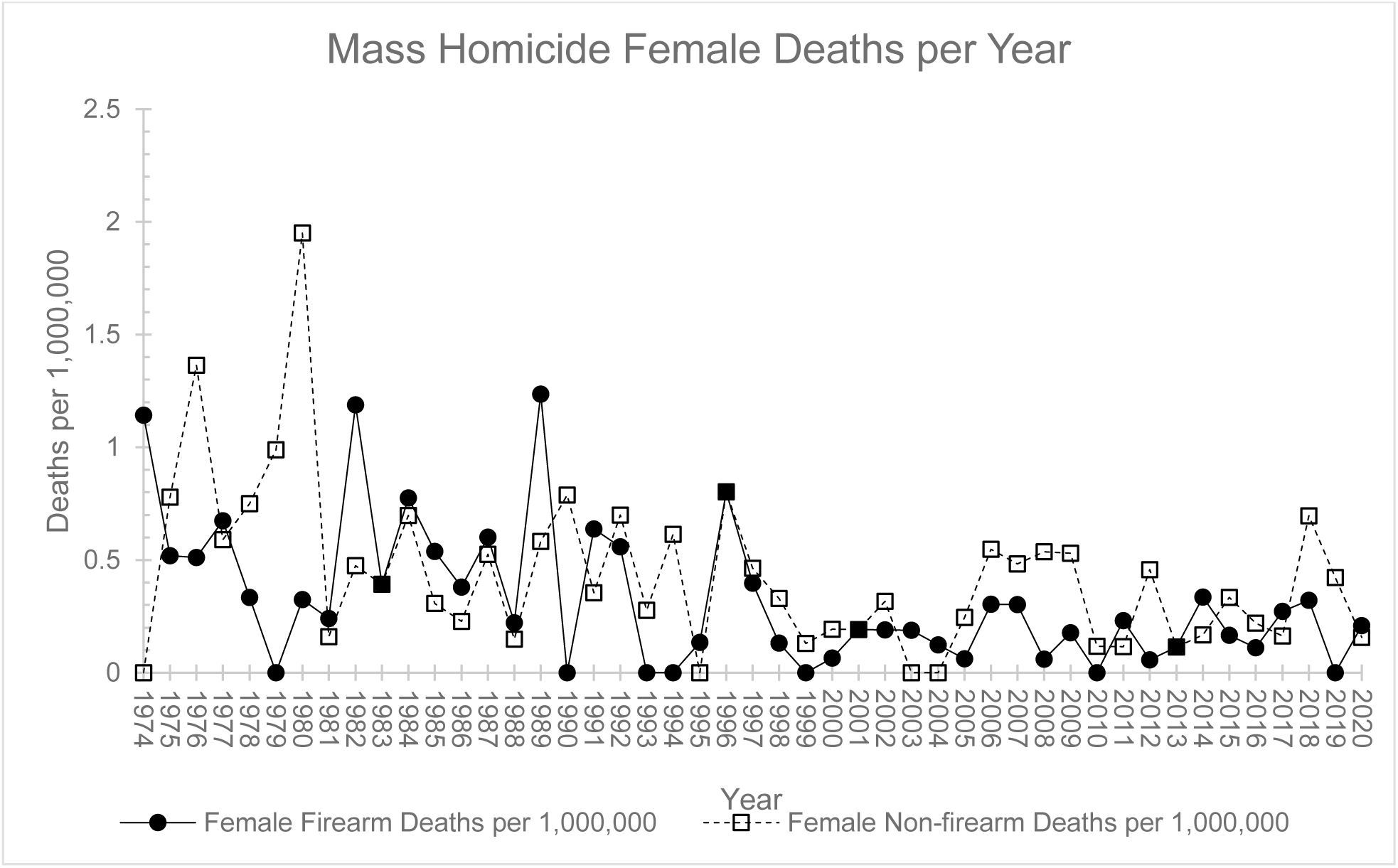

Table 1 contains descriptive statistics for the incidence rates of mass homicide by firearm and non-firearm as well as the death rates by weapon type and sex. The median number of mass firearm homicide incidents a year is 3, with an interquartile range (IQR) of 1 to 4, translating to a median of 11 (IQR 4, 16) deaths a year. The maximum number of incidents, 8, occurred in 1991 and 1992 while six years had no incidents at all. The maximum number of deaths, 33, occurred in 1982.

**Table 1.**
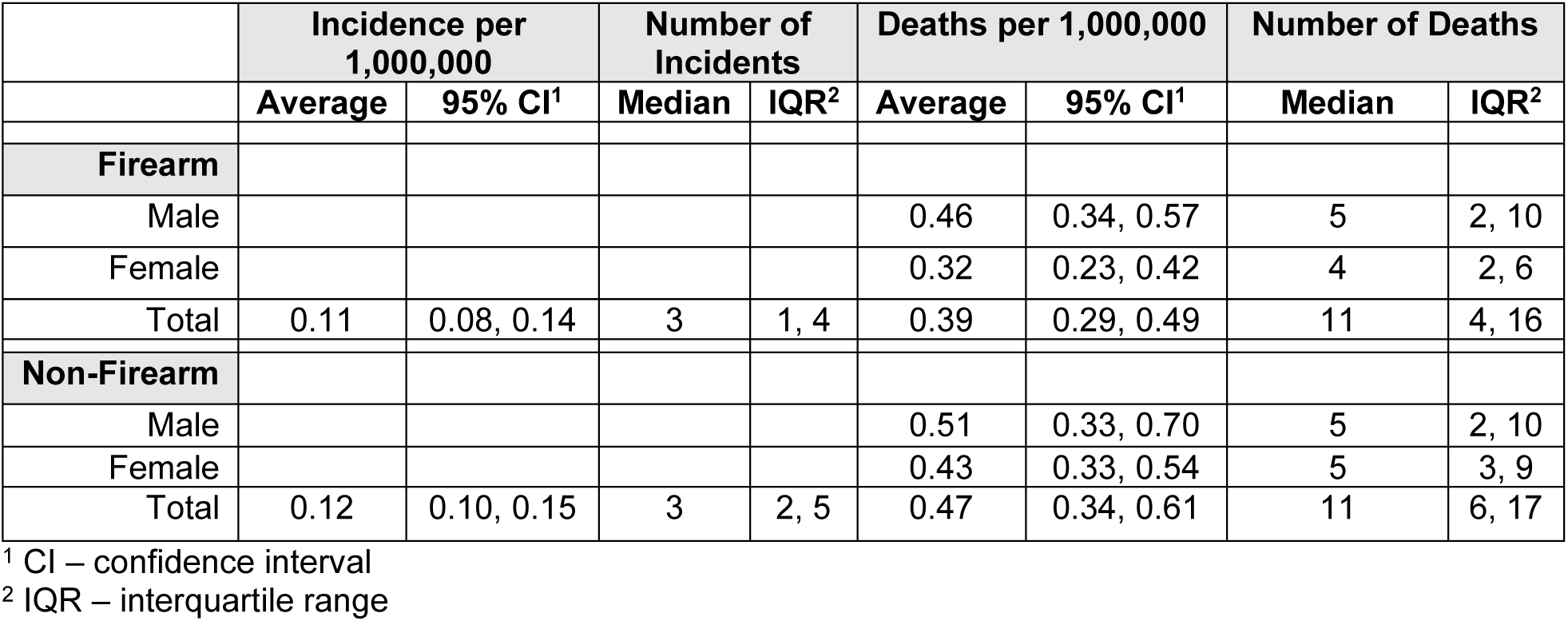
Incidence and Death Rates of Mass Homicide by Weapon Type and Sex.

Table 2 contains the results of the DiD analysis. The intervention in 1980 was associated with a significant IRR 1.57 (95% CI 1.02, 2.41) increase in the incidence of mass homicide by firearm compared to non-firearm. Linear combinations revealed that the incidence of firearm mass homicide did not change post the intervention, IRR 1.32 (95% CI 0.92, 1.91) per year, nor did the rate of non-firearm homicide, IRR 0.85 (95% CI 0.71, 1.01). Instead, a divergence between the rates after the 1980s appears to have occurred with mass homicide incidences by firearm increasing and mass homicide by non-firearm decreasing. The interventions in 1994 and 2001 did not have changes in incidences of mass homicide by firearm that were statistically different from the changes in incidents of mass homicide by non-firearm suggesting that the interventions had no association.

**Table 2.**
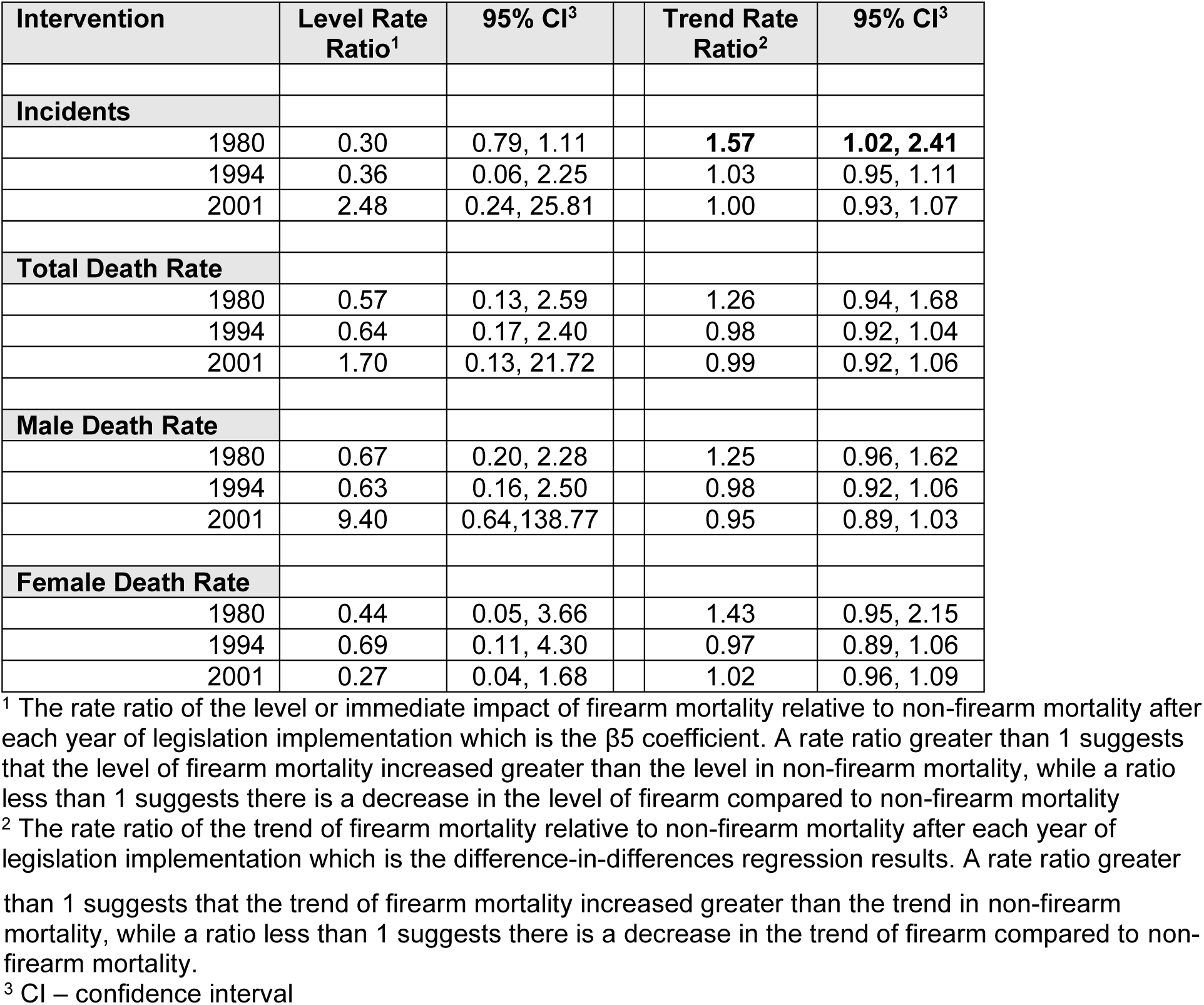
Incidence Rate Ratios of Firearm to Non-Firearm Post Intervention.

Total death rates by mass homicide did not have statistically different changes in rates compared to rates of mass homicide by non-firearm after the interventions in 1980, 1994 and 2001. Finally, death rates of mass homicide by firearm disaggregated by sex remained similar to death rates of mass homicide by non-firearm after the same interventions (Table 2).

## Discussion

This study found that, except after an intervention in the 1980s, the changes in the incidence of mass homicide were not statistically distinguishable before and after the implementation of legislation and regulations intended to decrease these occurrences such as background checks to purchase long guns, reference checks, psychological questionnaires, waiting periods, and licensing. After the banning of fully automatic firearms and the requirement of background checks to acquire long guns in the 1980s the incidence of mass homicide by firearm increased compared to the incidence of mass homicide by other methods. This may reflect a shift in behaviour by individuals intent on harming a large number of people or a change in criminal and gang behaviour (13, 14). Overall, however, the incidence of mass homicides gradually declined over the years 1974 to 2020.

There were, in addition, no statistically associated changes in death rates suggesting that the number of deaths associated with these incidents has not changed with implementation of legislation and regulations to decrease the lethality of firearms such as the prohibition of fully automatic firearms, prohibition of paramilitary style firearms, and magazine capacity limits. As well, the death rates by mass homicide gradually declined over the years 1974 to 2020.

The preponderance of recent studies regarding the association with legislation and mass homicide by firearms that use statistical controls and/or a time series analysis have been in the United States. The results of these studies are somewhat mixed and may suffer from the multiple comparisons problem resulting in false positives. Similar to this study, background checks and licensing for all types of firearms was not found to be associated with occurrences of mass shootings by Blau et al. (2016), while Webster et al. (2020) found only handgun licensing was associated with lower incidents and deaths (15, 16). Conversely Webster et al. (2020) did not find an increase in incidents in states that did not require a permit and found that permits were paradoxically associated with an increase in domestic mass homicide deaths. The associated benefit was lost when the cut off was increased from three to four victims. Siegel et al. (2020) found that permits were associated with a lower occurrence of a mass homicide incidents but not with the number of deaths, while background checks were not associated with any reductions (17).

Interestingly Siegal et al. (2020), and Webster et al. (2020), did not find any association with assault weapons bans and incidents of mass homicide and deaths in agreement with the findings in this study. Both studies, unlike this study, did find that there was a decrease in deaths associated with large capacity magazine bans, though for Webster et al. (2020) the results are not always consistent. When Webster et al. (2020) disaggregated the mass homicides into different categories, the reduction only remained associated with domestic violence related mass homicides. This dichotomy is difficult to understand as presumably a large capacity magazine would be more lethal in a mass public shooting. It is worthwhile to note that when Blau et al. (2016) examined mass homicides by specific weapon type, no one type of firearms was found to be more lethal than another.

Part of the difficulty in assessing firearm laws in the United States is the wide differences in firearm laws between states, allowing people to purchase a firearm in a less regulated state and bring it into a more regulated state. Canada provides a unique system to study firearm legislation as the firearms laws examined are created by the federal government and apply universally across provinces. In that way this study may be more robust in that mobility is not a confounding factor. This study also benefits from the ability to make causal links due to the DiD quasi-experimental design, whereas the Blau et al. (2016) and Siegel et al. (2020) studies are cross sectional studies. While the Webster et al. (2020) study is a time series study, unlike DiD studies, it may be more subject to issues due to confounders.

Limitations of this study include the inability to assess the types of firearms used in mass shootings, for example military style firearms, because data for this was not available. Only deaths were examined, no data was available for injuries occurring in the same event. As well data is unavailable to break down the mass shooting by location, whether it occurred in public such as work or schools, or at home, or both. Data about the perpetrator and motivations were unfortunately also not available. Only recently has data been collected on whether the accused had previous convictions (1997), whether they had a firearm license (2010), or it if the homicide was related to gang or criminal activity (1999) (18).

The most significant limitation is the small numbers of mass homicides and the fact that there are several years with no homicides that results in some cases of large confidence intervals and limited study power. Sensitivity testing revealed that for the interventions in the year 1994, by far the most significant type and number of interventions, an immediate level change in the incidence of firearm mass homicide resulting in 1.5 incidents less than the current number each year, would result in a statistically significant change compared to mass homicide by other methods. A decline in the trend of incidents by only 11% a year would also result in a statistically significant change. Therefore, it is possible to suggest, that an effective intervention would result in a detectable change.

## Conclusions

This is the first study to examine mass homicide in Canada as well as the association between legislative interventions controlling firearms and firearm mass homicide incidence rates and deaths. Over the period 1974 to 2020 the incidence and death rates associated with mass homicide gradually declined. Interestingly, interventions such as background checks, licensing, prohibition of military style firearms, and prohibiting large-capacity magazines, were not associated with changes in the incidence and deaths by mass homicide by firearms. The benefit from the consistency of firearms regulations in Canada eliminates the confounders in US studies due to the differing regulations across states. This study provides policy makers evidence to consider regarding future interventions to reduce deaths from firearm mass homicide. Recommended areas for data collection by Statistics Canada to aid future study include locations of incidents, the interpersonal relationship between perpetrator and victim, and the perpetrator’s motivation for mass homicide.

## Data Availability

All relevant data are within the manuscript and its Supporting Information files.

